# LUMANA: A Scenario-Based Field Validation of a Culturally Adapted AI System for Mental Health Screening in Sokoto State, Nigeria

**DOI:** 10.64898/2026.09.03.26362181

**Authors:** Tahir Buhari, Abubakar Baguda, Moses Aiyenuro, Jameel Ismail Ahmad, Jamil Galadanchi, Adnike Oluyori, Rufaida Musa Abdullahi, Zubairu Iliyasu, Ota Akhigbe, Atef Fawaz, Ruth Nkem, Aminu Ayuba, Hamisu Mohammed Salihu, Aisha Abdullahi Adam, Nafisat Amadu Abdulmalik

## Abstract

**Background:** Mental health conditions represent a significant public health burden in sub-Saharan Africa, where access to specialist services remains limited. Sokoto State, Nigeria, has an estimated population exceeding 5.4 million with fewer than five psychiatrists, leaving frontline health workers to provide much of the mental health care. LUMANA is a human-centred artificial intelligence (AI) system developed to support mental health screening in resource-constrained settings.

**Methods:** We conducted a field validation of LUMANA in Sokoto State, Nigeria, on 28–29 January 2026 following ethical approval (SKHREC/014/2026). Twenty stakeholders were enrolled, of whom 17 completed structured evaluations across eight standardized clinical scenarios using five assessment tools. The evaluation examined AI output safety, Hausa language performance, human–AI workflow integration, trust and cultural appropriateness, and implementation readiness.

**Results:** A total of 136 AI output safety assessments were completed. Overall, 113/136 (83%) AI outputs were rated acceptable as shown, while 23/136 (17%) were considered acceptable only with human review; no AI output was rated unacceptable. Religion and culture were identified as important determinants of trust by 14/17 (82%) participants, while 17/17 (100%) agreed that disclosures of self-harm or suicide should always involve a human clinician. Mean language quality scores were 9.9/10 for Hausa transcription, 9.9/10 for Hausa-to-English translation, and 9.2/10 for English-to-Hausa summary generation. Clinicians considered 7/8 (88%) human–AI workflow scenarios comfortable for routine practice. Three operational improvements were identified before pilot implementation: clearer protocols for interpreting distress, refinement of language to minimise potentially stigmatizing expressions, and automatic escalation of suspected psychotic symptoms.

**Conclusions:** This scenario-based field validation provides preliminary evidence that LUMANA is perceived as culturally appropriate, acceptable and capable of supporting human-supervised mental health screening in a low-resource setting. However, the findings are based on a small convenience sample using standardized scenarios rather than live clinical use. Accordingly, the results support progression to a carefully monitored pilot implementation following completion of the identified system improvements, with prospective evaluation of safety, workflow integration and implementation outcomes before consideration of wider deployment.

## 1. Introduction

### The Challenge: Mental Health in Low-Resource Settings

Mental health disorders affect approximately 1 billion people globally.^1^ However, the burden is distributed unequally, where low- and middle-income countries carry a disproportionate share of disease burden with minimal access to services.^2^ Moreover, depression and anxiety alone cost the global economy nearly $1 trillion annually in lost productivity.^3^ Nigeria alone is estimated to have approximately 40 million people living with mental health conditions.^4^ Yet, the mental health budget, mainly financed through the central government health budget, is about 3.3%–4%, with over 90 % going to the few neuropsychiatric hospitals available in Nigeria.^5^ With a population exceeding 220 million ^6^ and only about 250 psychiatrists serving the country, equivalent to approximately one psychiatrist per 800,000 people,^7^ the scarcity of specialist mental health professionals underscores the urgent need to strengthen mental health services. Consequently, the majority of Nigerians experiencing depression, anxiety, psychosis or other mental health conditions receive no formal care.

Sokoto State, located in northwest Nigeria, represents a typical setting in resource-limited sub-Saharan Africa. The state has an estimated population exceeding 5.4 million people.^8^ Psychiatric capacity is concentrated in the state capital. Rural communities where the majority of Sokoto’s population resides have virtually no mental health specialists.^9^ Instead, primary healthcare centers and community health volunteers serve as the frontline mental health workforce. These dedicated workers possess limited formal mental health training, minimal supervisory support and few tools to help them systematically identify mental health conditions in patients presenting with somatic complaints, behavioral changes or emotional distress.

### The Opportunity: AI as a Force Multiplier

Artificial intelligence offers genuine potential in this context, not as a replacement for clinical judgment but as a force multiplier. An AI system that could help frontline health workers recognize mental health symptoms in patients’ own language, respect cultural and religious frameworks for understanding psychological distress, and know when to escalate to specialists could meaningfully improve mental health case identification and access to care.

However, a persistent limitation of AI in mental health is that training datasets rarely reflect the full spectrum of cultural backgrounds, languages and socio-demographic profiles found in real-world populations, creating risk of entrenching existing health inequities or generating new forms of algorithmic bias, particularly at the points of diagnosis and triage.^10^ Many public health AI models, however, draw from data sets in populations which are unrepresentative of those in the low- and middle-income countries (LMICs). The resulting data inequity means the algorithms then do not capture the cultural, linguistic, genetic, or environmental variety in the underserved populations.^11^ Hence, LUMANA was designed differently.

### LUMANA: Design Philosophy

LUMANA represents a deliberate pivot toward AI systems designed specifically for resource-constrained, culturally diverse settings. The system was built from its inception with three non-negotiable design principles.

First, human-centered design. The system exists to support human decision-making, not replace it. In high-risk scenarios, such as suicidal ideation, psychotic symptoms, severe emotional distress and language and trust breakdown, the AI’s role is strictly limited to detection and escalation. All clinical judgment remains with the clinician. The system is transparent; clinicians can see its reasoning and can override its recommendations when appropriate.

Second, cultural competence is a mandatory requirement. Mental health is deeply culturally embedded. How people in Sokoto understand and express psychological distress differs fundamentally from how people in Western contexts describe their struggles. Idioms of distress are a well-documented, globally recurring way that psychological suffering is communicated through locally meaningful language rather than biomedical symptom terms.^12^ A system that ignores this reality would be both ineffective and harmful. LUMANA was trained on Hausa language data, tested with Hausa speakers and evaluated by clinicians familiar with how mental health presents in northwestern Nigeria. The system understands Hausa idioms of distress and respects Islamic and Hausa cultural values.

Third, transparency and safety are prioritized over sophistication. The team resisted pressure to build a system that made impressive predictions but couldn’t explain its reasoning. Instead, LUMANA generates assessments that clinicians can read, understand and reason about. When the system is uncertain, it says so. When human judgment should override the system, clinicians are empowered to do so.

### Research Questions

This field validation addressed a central question: Is LUMANA safe, culturally appropriate and acceptable for use by frontline health workers in mental health screening in Sokoto State? This overarching question encompasses several specific sub-questions:

1. Can LUMANA accurately identify mental health symptoms when patients describe them in Hausa, their first language?
2. Does the system generate responses that are culturally appropriate and respectful of Islamic and Hausa cultural frameworks?
3. Will clinicians trust the system enough to incorporate it into their workflows?
4. Are there specific high-risk scenarios where the system should not attempt clinical judgment?
5. What operational infrastructure is required before patient-facing deployment can begin?

## 2. Methods

### Ethical Approval and Regulatory Framework

This field validation was conducted under full ethical oversight from the Sokoto State Health Research Ethics Committee (SKHREC). The protocol titled “Conduct of Lived Experience Engagement Activity at Gandi Rabah IDP Camps, Sokoto State” received approval on 26 January 2026 with reference number SKHREC/014/2026. Approval was granted by Bashiru Bello, Director of Health Planning, Research and Statistics, Sokoto State Ministry of Health.

All study procedures adhered to the principles expressed in the Declaration of Helsinki. All participants provided documented informed consent for voluntary participation, anonymized data use, optional audio recording and acknowledged their right to withdraw at any time without consequence. Clinicians were available throughout the validation for participant support.

### Study Design and Setting

The validation employed a mixed-methods design combining qualitative clinical scenario evaluation with quantitative participant feedback. The qualitative components of the study were reported in accordance with the Standards for Reporting Qualitative Research (SRQR) to ensure transparent and comprehensive reporting. The study was conducted in Sokoto State, Nigeria, on 28-29 January 2026. This two-day intensive field validation was conducted in a setting representative of sub-Saharan African contexts: significant mental health burden, limited specialist capacity, predominantly Hausa-speaking population, substantial internally displaced person (IDP) populations and frontline health workers as the primary mental health workforce.

### Participant Population

Twenty participants were enrolled in the validation. Seventeen participants provided structured feedback via validated data collection tools. The participant composition reflected diverse stakeholder groups:

1. Lived experience participants (n=12, 60%). Individuals with direct personal or family experience of mental health conditions. All were adults aged 18 years or older, meeting ethical requirements for informed consent.
2. Clinicians (n=5, 25%). Experienced primary care providers, mental health nurses and one psychiatrist from the Federal Neuro-Psychiatric Hospital Kware in Sokoto.
3. Community health worker (n=1, 5%). One peer counselor/community health worker represents the frontline workers who would ultimately use LUMANA in clinical practice.
4. Project staff (n=2, 10%). Project leadership and eHealth Africa team members.

All participants consented individually and confirmed willingness to have anonymized data used for research and reporting.

### Sampling strategy

The sample was drawn through a non-random convenience sampling strategy to ensure representation across the population. Recruitment targeted three stakeholder categories relevant to LUMANA’s intended use, including lived experience individuals, clinicians and community health workers

### Data Collection Tools

Five structured data collection tools were administered:

- Participant Consent & Safeguarding Form (Tool 1): This form documented informed consent across all required domains. All participants provided full consent.
- Trust & Acceptability Reflection (Tool 2): This open-ended questionnaire asked participants to identify situations where they would trust an AI-supported mental health system and situations where they would not. Participants identified “red lines”—absolute boundaries the system must never cross.
- AI Output Safety Review (Tool 3): Administered to 17 participants (12 lived experience and 5 clinicians), this was the primary evaluation tool. Each participant reviewed eight mental health scenarios and corresponding LUMANA outputs, indicating whether outputs were acceptable as shown or acceptable only with human review.
- Hausa Idioms of Distress Mapping(Tool 4): Language experts and bilingual clinicians reviewed four Hausa expressions of emotional distress commonly used in Sokoto, classified them by frequency of use, emotional states they typically express and demographic patterns.
- Human-AI Workflow Evaluation (Tool 5): This tool asked clinicians to specify appropriate AI role, human role, risk level, escalation requirements and key recommendations for each scenario.^13^

### Clinical Scenarios

The validation tested LUMANA across eight realistic mental health presentations:

- Mild anxiety: Somatic symptoms (heat in body, heart pounding, sense of collapse)
- Moderate depression: Persistent low mood, hopelessness, sleep problems, fatigue (3-month duration)
- Suicidal ideation: Passive death wishes (“Sometimes I think death is better for me”)
- Psychotic symptoms: Paranoia and persecutory ideation
- Ambiguous distress: Somatic symptoms attributed to “spiritual imbalance and social pressure”
- Repeat screening and trend tracking: Depression worsening over 3 months (PHQ-9: 14→18) ^14^
- Language and trust breakdown: System generates stigmatizing or offensive language
- Clinician disagreement: Clinician overrides system recommendation based on clinical judgment

These scenarios were not hypothetical edge cases but realistic presentations regularly encountered in Sokoto that pose genuine clinical challenges for frontline health workers.

### Data Analysis

#### Quantitative analysis

For the AI Output Safety Review form (17 participants × 8 scenarios = 136 reviews), we calculated frequencies and percentages for categorical variables. Descriptive statistics were generated for tone appropriateness. Language distribution was analyzed by AI Output ID.

#### Qualitative analysis

Processing of audio recordings included transcription and translation from Hausa to English, and each transcript was reviewed for translation accuracy and vocabulary appropriateness. Participant identifiers were removed and responses were anonymised prior to analysis.

Clinician narrative feedback was analyzed thematically to identify consensus points, areas of concern and scenario-specific recommendations. Hausa idioms documentation was analyzed by emotional context, demographic use patterns and safety classification.

#### Synthesis analysis

Findings across the five data collection tools were triangulated to identify convergent evidence across different stakeholder perspectives.

### Researcher Characteristics and Reflexivity

The validation was designed and conducted by a multidisciplinary team combining expertise in digital health, clinical psychiatry, community medicine, epidemiology, and Hausa language and culture. The team included 3 facilitators present during the sessions, clinicians drawn from primary care, mental-health nursing and psychiatry (including a psychiatrist from Federal Neuro-Psychiatric Hospital Kware, Sokoto), academic collaborators in public health and psychiatry from the Kano Independent Research Centre Trust, and language and cultural experts native to northwest Nigeria. The majority of the team are Nigerian and Hausa-speaking, with direct familiarity with how psychological distress is expressed and understood in Sokoto. This shared linguistic and cultural background was an asset for interpreting idioms of distress and assessing cultural appropriateness, but the team was also conscious that it could predispose evaluators toward favourable interpretation, and this was weighed during analysis.

Several members of the team, including the corresponding author, are affiliated with eHealth Africa, the organisation that developed LUMANA and funded the validation through a Wellcome Trust grant. The team was therefore invested in the system’s success, which constitutes a potential source of confirmation and social-desirability bias (also noted in the Limitations). To mitigate this, facilitators emphasised to participants that candid and critical feedback was the primary purpose of the exercise; responses were anonymised and recorded under participant codes (P01–P17); clinical scoring was performed by clinicians who were not part of the LUMANA development team; and no member of the development team scored the AI outputs. Clinical oversight throughout was provided by Dr Nirmal Ravi (Chief Innovations Officer, eHealth Africa), who was independent of day-to-day system development.

The team approached the validation with the prior assumption that culturally grounded design would matter to acceptability and safety; this assumption shaped the choice of tools (e.g the idioms-of-distress mapping) and should be borne in mind when interpreting the strongly positive cultural-appropriateness findings. No formal member-checking of interpretations with participants was undertaken, and inter-rater reliability for the language scores was not assessed, both of which temper the strength of the qualitative claims.

## 3. Results

### 3.1 Overview

Seventeen participants completed structured validation across eight standardized mental health scenarios, generating 136 AI output safety assessments. Of these, 113 (83%) were rated as acceptable as presented, while 23 (17 %) required human review before use. No scenario received a majority rating of either unacceptable or unsafe, indicating overall acceptable system performance during scenario-based evaluation.

### 3.2 Trust and Acceptability Reflection

Participants generally trust the AI-supported mental health system in situations where human support is limited, such as in IDP camps or areas with no nearby mental health facilities, for purposes such as mental health education, personal assessments, emotional tracking, and reducing clinician workload.

Participants indicated that they would not trust or would stop using the AI-supported mental health system in situations involving mental health emergencies, psychiatric crises, or high-risk scenarios where professional human support, such as a psychiatrist or psychologist, is available.

Religion and culture were identified as the primary trust boundary, with 82% (14/17) of participants identifying these as a red line that should not be crossed, while 12% identified marital issues and 6% did not specify.

### 3.3 AI Output Safety Assessment

A total of 12 lived experience participants and 5 clinicians were administered the form, giving a total of 17 participants. The language distribution of the reviews was Hausa and English combined (91%), Hausa only (7%), and English only (1%). Across all scenarios, the majority of participants reported feeling supported and understood by the AI outputs, and most tone assessments were rated as appropriate either without restriction or with human oversight.

The table below presents the overall safety assessment of the AI tool across eight mental health scenarios (HC-01 to HC-08). Participants were asked to indicate whether the AI output was acceptable as shown or acceptable only with human review.

Overall safety assessment of LUMANA AI outputs is presented in Table 1. A total of 136 AI-generated responses (17 responses across each of the eight scenarios) were independently assessed. Of these, 113/136 (83%) were rated as acceptable as shown, while 23/136 (17%) were considered acceptable only with human review. No AI output was rated as unacceptable or not allowed for use. At the scenario level, the proportion of outputs judged acceptable as shown ranged from 11/17 (65%) for HC-01 to 17/17 (100%) for HC-03. All outputs for HC-05, HC-07 and HC-08 were considered acceptable as shown or acceptable with human review, with no unacceptable ratings recorded. HC-01 required the greatest level of human oversight, with 6/17 (35%) outputs judged acceptable only with human review, followed by HC-02 (4/17; 24%) and HC-06, HC-07 and HC-08 (3/17; 18% each).

**Table 1:** Overall safety assessment of LUMANA AI outputs by scenario.

| Overall Safety Assessment | HC-01 | HC-02 | HC-03 | HC-04 | HC-05 | HC-06 | HC-07 | HC-08 | Total |
| --- | --- | --- | --- | --- | --- | --- | --- | --- | --- |
| Acceptable as shown | 11 | 13 | 17 | 15 | 15 | 14 | 14 | 14 | 113 |
| Acceptable only with human review | 6 | 4 | 0 | 2 | 2 | 3 | 3 | 3 | 23 |
| **Total** | **17** | **17** | **17** | **17** | **17** | **17** | **17** | **17** | **136** |

**Source: Research survey**

**Table 2:** Human–AI workflow evaluation by scenario.

| Scenario | Risk Level | Escalation Required | Clinician Comfort |
| --- | --- | --- | --- |
| HC-01: Mild Anxiety | Low–Medium | No | Comfortable |
| HC-02: Moderate Depression | Low | Conditional(PHQ-9 threshold) | Comfortable |
| HC-03: Suicidal Ideation | High | Yes | Comfortable |
| HC-04: Psychotic Symptoms | High | Yes | Comfortable |
| HC-05: Ambiguous Distress | High | Yes | Comfortable |
| HC-06: Repeat Screening / Follow-up | High | Conditional if the score goes beyond moderate | Comfortable |
| HC-07: Language & Trust Breakdown | High | Yes | Somewhat uncomfortable |
| HC-08: System Recommendation Disagreement | Medium | Yes | Comfortable |

**Source: Authors' computation**

Overall, these findings indicate that all LUMANA AI outputs were considered suitable for use, either without modification (83%) or with human oversight (17%). The need for human review was concentrated in a small number of scenarios, particularly HC-01, suggesting that although the system demonstrated a favourable safety profile overall, clinician oversight remains important for selected clinical situations.

### 3.4 Hausa Language Quality and Translation Accuracy

Evaluators scored each translation stage on a 0–10 scale, with remarkably strong results:

Hausa-to-Hausa transcription averaged 9.9/10, Hausa-to-English translation averaged 9.9/10, and English-to-Hausa mental health summary translation averaged 9.2/10. Word error rates ranged from 0 to 2 across the 8 scenarios, which is clinically acceptable.

Overall translation performance across the three language-processing stages (Hausa-to-Hausa transcription, Hausa-to-English translation, and English-to-Hausa summary generation) yielded a mean score of approximately 9.6/10, indicating consistently high language accuracy during the validation exercise. This summary measure reflects the average of the three stage-specific mean scores (9.9, 9.9 and 9.2) reported above and provides an overall indicator of language performance across the Hausa–English–Hausa workflow.

Beyond technical accuracy, the validation also examined whether LUMANA understood and appropriately used Hausa idioms of distress. Language experts compiled a bank of culturally specific expressions, each classified for its safety in AI processing.

Of these, “Akwai rina a kaba” (there is something hidden) is used to signal that a person is anticipating something negative. This expression is classified as unsafe for AI recognition, as it carries a risk of mislabelling and requires the system to probe for additional detail before concluding.

The remaining three expressions were classified as requiring careful interpretation rather than being inherently unsafe: “Zuciya ta tanada nauyi” (my heart feels heavy), which can signal sadness, hopelessness, anxiety, or worry; “Ina cikin yanayi (matsala)” (I am in a particular situation), which signals broader emotional distress; and “Kai dai a yi sha’ani,” which marks a deep issue the person is not yet ready to disclose. Each of these may lead the AI to mislabel, over- or underestimate distress if interpreted literally, and therefore also requires probing for additional detail.

The creation and documentation of this Hausa idioms-of-distress bank represents a meaningful contribution to AI-assisted mental health work, demonstrating that cultural competence requires deliberate, careful effort to understand how distress is expressed within specific languages and cultures.

### 3.5 Human–AI Workflow Evaluation

Clinicians rated seven of eight scenarios (87.5%) as comfortable and one scenario (12.5%), Language and Trust Breakdown (HC-07), as somewhat uncomfortable. Five scenarios (HC-03, HC-04, HC-05, HC-06 and HC-07) were classified as high risk, (HC-08) as low-to-medium risk (HC-01) and (HC-02) as Low. Escalation was required in all high-risk scenarios and was conditional for moderate depression and repeat screening.

## 4. Deployment recommendation

### Overall Recommendation

After comprehensive field validation involving 20 stakeholders across 8 clinical scenarios, with data triangulation across clinical judgment, quantitative safety assessment, and community perspectives, we recommend proceeding with patient-facing deployment of LUMANA in Sokoto State, contingent upon full completion of the three critical gaps identified in the Results section.

This recommendation is based on substantial evidence of system safety and effectiveness:

1. Clinical Signal Detection: The system correctly identifies mental health symptoms across diverse presentations and appropriately stratifies risk.
2. Language Quality: As demonstrated in the Results (Section 3.4), LUMANA achieved an overall mean translation performance of approximately 9.6/10 across the Hausa–English–Hausa workflow, with high scores for transcription, translation and summary generation. In addition, the system appropriately recognised Hausa idioms of distress and generated culturally appropriate language, supporting its potential suitability for clinical screening with appropriate human oversight. Clinician Trust: Seven of eight scenarios were rated “Comfortable” or “Very Comfortable” by clinicians. Only one scenario with lower comfort ratings corresponds directly to the three critical gaps, which are resolvable through specific mitigation strategies.
3. Community Acceptability: Eighty-three percent of AI outputs were rated as acceptable as shown by 17 stakeholders, including patients and clinicians. Eighty-one percent reported no identified issues. Overwhelmingly, participants reported feeling “Supported & Understood” after reading system outputs.
4. Human-Centered Design: The system appropriately restricts its role to detection and escalation for high-risk cases, with human clinicians making all critical clinical judgments.
5. Cultural Respect: The system demonstrates commitment to respecting Islamic faith and Hausa cultural values, with documented protocols for cultural interpretation.

However, this recommendation comes with a clear conditional statement: the three critical gaps must be closed before patient-facing deployment. Responsible deployment of AI in healthcare requires acknowledging both what is working and what still needs to be done.

### Phased Deployment Pathway

The recommended deployment pathway is:

### Phase 0: Gap Closure (26 January – 28 February 2026)

Complete all three critical gap-closure activities. No patient-facing deployment occurs during this phase. By the end of February, the team should demonstrate: (1) language audit completion with documentation of all revisions, (2) clinician training curriculum finalized and trainers prepared, (3) escalation protocols documented and mock drills completed, (4) system architecture modifications implemented and tested.

### Phase 1: Pilot Deployment (1 March – 30 April 2027)

Launch LUMANA pilot at 2-3 health facilities in Sokoto with strong leadership commitment, reliable clinician staffing and good technical infrastructure. Deliver full 40-hour clinician training at each site. Establish daily monitoring (one designated person reviewing escalations, technical issues, language problems and clinician feedback). During this phase, collect data on patient volume, escalation frequency and success, clinician satisfaction, technical issues and patient feedback.

### Phase 2: Expanded Deployment (May – December 2027)

If Phase 1 pilot demonstrates safety and acceptability, expand to 8-10 additional health facilities across Sokoto. Begin quarterly refresher training. Transition from daily monitoring to weekly review. Establish a sustainability plan.

### Phase 3: Full Implementation (2028)

Scale to all interested health facilities across Sokoto with demonstrated demand and readiness. By the end of 2028, the aim is to have LUMANA available in at least 20 health facilities serving approximately 50,000 patients annually.

## 5. Discussion

### 5.1 Culturally-Centered AI Design as Necessity, Not Choice

The findings highlight the importance of culturally informed AI design in mental health screening. LUMANA was not designed by starting with a general-purpose AI system and then translating it into Hausa. Rather, it was designed from the ground up with Hausa speakers, cultural experts and clinicians from Sokoto involved in every stage. The high language validation scores and strong acceptability ratings observed in this study suggest that effective mental health technologies require more than linguistic translation alone; they must also account for local cultural meanings and expressions of distress. The mental health experiences of people in Sokoto are genuinely different from those of people in Boston or Berlin. These differences are not superficial—requiring only translation—but fundamental, requiring deep cultural understanding embedded at every level of system design.

This observation is consistent with the finding that cultural adaptation makes interventions more relatable to the new target group by considering their specific context, burdens and understanding of mental health.^15^ Furthermore, cultural concepts and idioms of distress are increasingly recognized as important mechanisms through which emotional suffering is communicated and understood across societies.^16^ The identification of Hausa idioms of distress during validation supports the importance of incorporating culturally grounded expressions into AI-assisted mental health screening tools.

The validation’s identification of religion and culture as non-negotiable red lines cited by 82 % of respondents is not a limitation imposed on the technology but an affirmation by community members that mental health care in Sokoto must honor Islamic faith and Hausa cultural values. Previous findings highlighted the importance of ensuring that digital mental health systems are aligned with local cultural and religious values and are responsive to the communities they serve. Cultural adaptation is a key means of increasing the engagement of target communities.^17^ The Hausa idioms documented in this validation represent a beginning, not a complete inventory. Contextual interpretation of these expressions requires deliberate clinical and cultural expertise that cannot be embedded in AI systems through translation alone.

### 5.2 Safety Performance and Human Oversight

The safety evaluation demonstrated generally positive performance across all eight validation scenarios. Despite these positive findings, participants consistently emphasized the importance of human oversight, particularly in situations involving self-harm, suicide risk, psychotic symptoms, or culturally complex presentations. All participants agreed that disclosures involving self-harm or suicide require human involvement. The Language and Trust Breakdown scenario generated the lowest clinician comfort rating and was the only scenario classified as somewhat uncomfortable. These findings are consistent with the WHO guidance that AI systems used in healthcare should remain under human supervision, with humans retaining control over healthcare systems and medical decisions. ^18^ Maintaining trust is particularly important in mental health settings, where communication quality directly influences engagement and disclosure. These findings indicate favourable stakeholder perceptions of safety and cultural appropriateness under standardized scenario conditions, but they do not establish clinical effectiveness or diagnostic accuracy. Prospective evaluation with real-world patient interactions and independent outcome verification is still required.

### 5.3 Workflow Integration

The workflow evaluation confirms that LUMANA is most safely deployed as a support tool in a tiered system: fully automated outputs are appropriate for low-risk cases; conditional or human-led responses are required for moderate cases; and human override with immediate escalation is non-negotiable for high-risk scenarios. The clinician comfort rating of 7/8 scenarios as ‘Comfortable’ suggests that the proposed workflow is acceptable to the clinical staff who would implement it. This pattern of acceptable performance and clinician comfort in 87.5% of scenarios supports progression to a carefully monitored pilot implementation rather than immediate routine deployment.

### 5.4 Limitation of the study

This study has several limitations. The validation used a small, non-random convenience sample (20 enrolled; 17 completed structured evaluation) without a comparison group. Participants assessed pre-prepared scenario outputs rather than interacting with a live LUMANA system, and no operational software was deployed during the validation. Because some investigators were affiliated with the system developer, social-desirability and evaluator-presence bias cannot be excluded. Inter-rater reliability for the 0–10 language ratings was not formally assessed. In addition, the study was conducted over two days in a single state and primarily within one ethnolinguistic context (Hausa-speaking populations), which limits generalisability. Future studies should prospectively evaluate LUMANA in routine clinical settings using larger, independent, multi-site samples.

### 5.5 What This Validation Reveals About Mental Health in Sokoto

First, there is genuine clinical competence and commitment among Sokoto’s frontline health workers. The clinicians and community health workers who participated brought a sophisticated understanding of how mental health presents in their communities. This competence and commitment are an enormous asset.

Second, the mental health gap in Sokoto is not primarily a knowledge problem; it is a capacity and resource problem. Frontline health workers know roughly what to do when they encounter mental health conditions. The barrier is that they are overwhelmed with patients, lacking supervisory support and working in settings with virtually no psychiatric backup. LUMANA’s value is that it frees them to apply the clinical knowledge they already possess.

Third, culture and religion are not obstacles to mental health care in Sokoto; they are foundations upon which effective mental health care must be built. The identification of “religion and culture” as a non-negotiable red line was an affirmation by community members that mental health care in Sokoto must honor Islamic faith and Hausa cultural values.

## 6. Conclusion

This scenario-based field validation provides preliminary evidence that LUMANA is perceived as culturally appropriate, acceptable and potentially useful for supporting mental health screening within a human-supervised care model in Sokoto State. However, given the short duration, small convenience sample and non-live evaluation design, the findings support progression to a monitored pilot implementation following completion of the identified operational improvements, rather than a conclusion that the system is ready for routine deployment.

The evidence supporting this conclusion is substantial. The system demonstrates excellent translation accuracy in Hausa (9.6/10), appropriate risk stratification of mental health presentations (correctly identifying 62.5% as high risk), strong cultural alignment with how mental health is understood locally and community acceptability (83% of outputs rated as appropriate as shown). Clinicians expressed high comfort with the system when decision boundaries were clear. Community members affirmed that the system respects religious and cultural values.

However, this positive conclusion comes with an essential condition: three specific, operationally defined gaps must be closed before patient-facing deployment. These gaps are not evidence of fundamental system failure. Rather, they are concrete issues with clear solutions that can be completed within four weeks.

Mental health remains profoundly underaddressed in sub-Saharan Africa. In Sokoto State, with over 5.4 million people and fewer than five psychiatrists, the vast majority of people experiencing depression, anxiety, psychosis, or other mental health conditions receive no formal care. LUMANA cannot solve this crisis alone. But a well-designed, culturally-centered AI system that helps frontline health workers recognize mental health symptoms, that escalates appropriately for emergencies, and that respects community values could meaningfully improve mental health case identification and access to care for tens of thousands of people.

If LUMANA is deployed successfully in Sokoto—if frontline health workers feel supported rather than monitored, if patients feel understood and respected, if mental health case identification increases, if more people access the care they need—it will be because the system respected these boundaries and maintained humans as essential, irreplaceable decision-makers.

## Data Availability

### Underlying dataset

LUMANA Field Validation Dataset, Sokoto 2026 ^19^

Available at: https://doi.org/10.6084/m9.figshare.33204894

This project contains the following underlying data, all of which were collected during the 28–29 January 2026 field validation:

- Dataset 1: Lumana_Trust_Acceptability_Reflection_Dataset.xlsx — 17 participant responses on trust, acceptability, and “red lines” (Tool 2).
- Dataset 2: Lumana_AI_Output_Safety_Review_Dataset.xlsx — 136 valid reviews (17 reviewers × 8 AI outputs, IDs HC-01 to HC-08), per-output safety assessment, tone appropriateness, escalation triggers (Tool 3).
- Dataset 3: Lumana_Hausa_Idioms_of_Distress_Mapping_Dataset.xlsx — clinician-reviewed Hausa idioms of distress with safety classification (Tool 4).
- Dataset 4: Lumana_Human_AI_Workflow_Evaluation_Dataset.xlsx — clinician workflow appropriateness ratings per scenario (Tool 5). Participant identifiers in all datasets have been re-coded (P01–P17) and any free-text fields screened for indirect identifiers before deposit. Data are available under the terms of the Creative Commons Attribution 4.0 International license (CC-BY 4.0).

### Extended data

This contains LUMANA Data Collection instruments and supplementary material: (https://doi.org/10.6084/m9.figshare.33183916)

This collection contains the five data-collection instruments used in the LUMANA field validation study, provided in blank form and in English:

- Tool 1: Participant Consent & Safeguarding Form.
- Tool 2: Trust & Acceptability Reflection Form.
- Tool 3: AI Output Safety Review Form.
- Tool 4: Hausa Idioms of Distress Mapping Form
- Tool 5: AI Clinical Workflow Evaluation ODK Form. Data are available under the terms of the Creative Commons Attribution 4.0 International license (CC-BY 4.0)

**Supplementary Material:** Standards for Reporting Qualitative Research (SRQR) checklist.

## Software Availability

This study evaluated LUMANA using a scenario-based methodology: participants reviewed and assessed eight pre-prepared, clinically realistic AI outputs through structured surveys and interviews, rather than interacting with a deployed or live system. No software was run by or made accessible to participants during this validation.

## Author Contributions (CRediT)

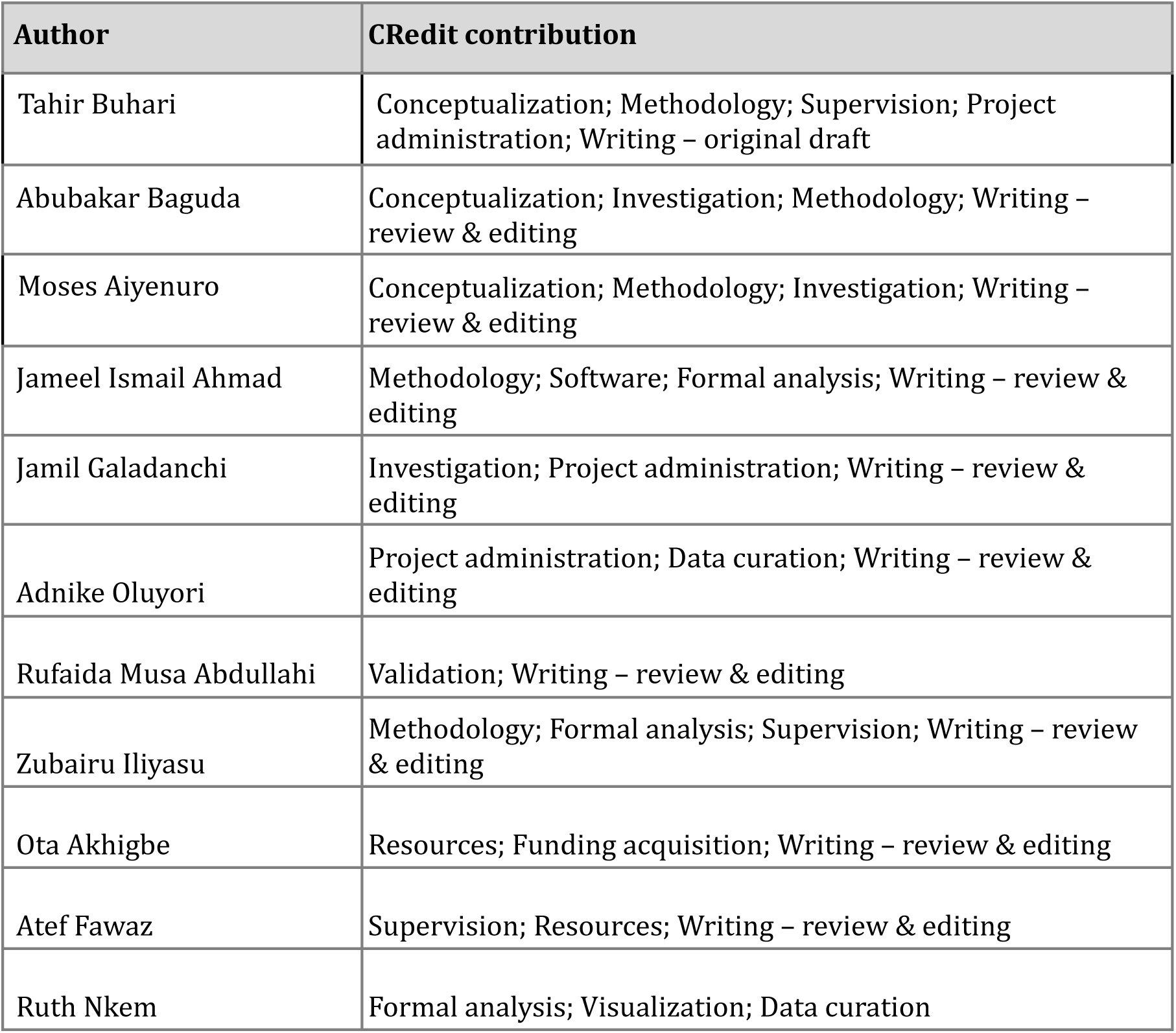
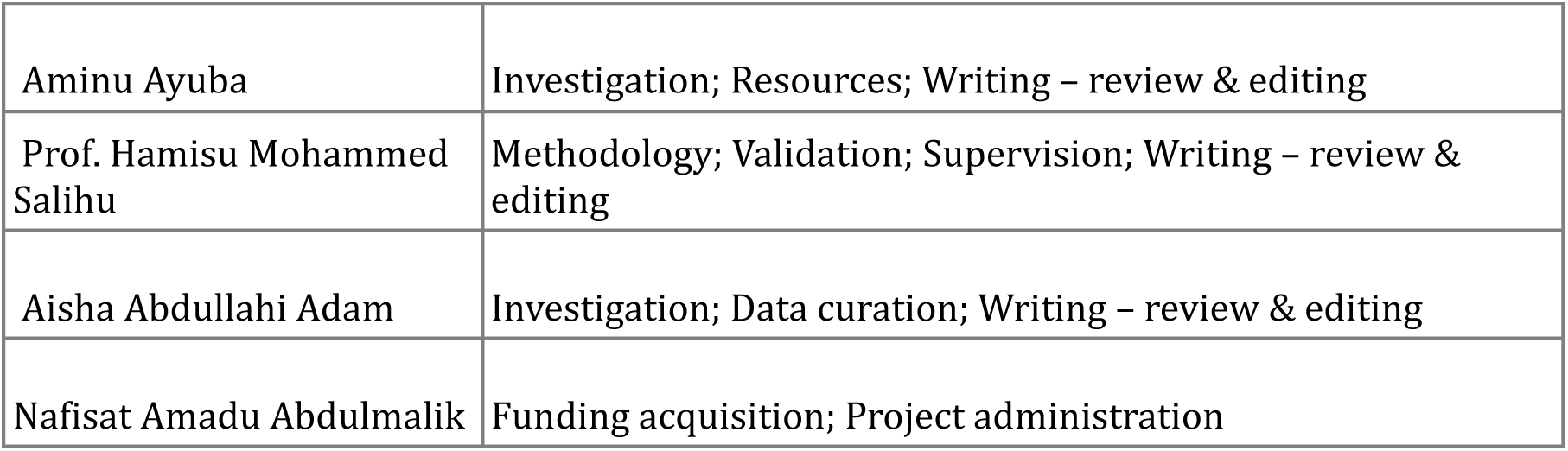

| Author | CRedit contribution |
| --- | --- |
| Tahir Buhari | Conceptualization; Methodology; Supervision; Project administration; Writing – original draft |
| Abubakar Baguda | Conceptualization; Investigation; Methodology; Writing – review & editing |
| Moses Aiyenuro | Conceptualization; Methodology; Investigation; Writing – review & editing |
| Jameel Ismail Ahmad | Methodology; Software; Formal analysis; Writing – review & editing |
| Jamil Galadanchi | Investigation; Project administration; Writing – review & editing |
| Adnike Oluyori | Project administration; Data curation; Writing – review & editing |
| Rufaida Musa Abdullahi | Validation; Writing – review & editing |
| Zubairu Iliyasu | Methodology; Formal analysis; Supervision; Writing – review & editing |
| Ota Akhigbe | Resources; Funding acquisition; Writing – review & editing |
| Atef Fawaz | Supervision; Resources; Writing – review & editing |
| Ruth Nkem | Formal analysis; Visualization; Data curation |
| Aminu Ayuba | Investigation; Resources; Writing – review & editing |
| Prof. Hamisu Mohammed Salihu | Methodology; Validation; Supervision; Writing – review & editing |
| Aisha Abdullahi Adam | Investigation; Data curation; Writing – review & editing |
| Nafisat Amadu Abdulmalik | Funding acquisition; Project administration |

## Competing interests

Several authors are employees of, or affiliated with, eHealth Africa, the organisation that developed LUMANA and provided financial and technical support for this validation. The corresponding author is an eHealth Africa research lead, and the organisation’s Chief Innovations Officer provided clinical oversight. These relationships constitute potential competing interests. The Wellcome Trust had no role in the study design, data analysis, interpretation of findings, manuscript preparation, or the decision to submit the manuscript for publication.

## Funding information

This research was supported by the Wellcome Trust [332492/Z/25/Z]. The funder had no role in study design, data collection and analysis, decision to publish, or preparation of the manuscript.

## Acknowledgements

This field validation was conducted with the financial and technical support of eHealth Africa, with ethical oversight from the Sokoto State Health Research Ethics Committee. We acknowledge Bashiru Bello (Director of Health Planning, Research and Statistics, Sokoto State Ministry of Health) for ethical approval and support; Dr. Nirmal Ravi (Chief Innovations Officer, eHealth Africa) for clinical oversight; and Hausa language experts who documented idioms of distress. We particularly thank the 20 community members, clinicians, community health workers and lived experience participants who generously gave their time and insights during this validation and the many community members in Sokoto who shaped the development and validation of LUMANA.

## Notes

### Author Declarations

Sokoto State Health Research Ethics Committee (SKHREC), Sokoto State Ministry of Health, Nigeria, gave ethical approval for this work (protocol "Conduct of Lived Experience Engagement Activity at Gandi Rabah IDP Camps, Sokoto State," reference SKHREC/014/2026, approved 26 January 2026).

